# Analysis of the Time-Dependent Behaviors of Atrial Fibrillation with Electrographic Flow Mapping

**DOI:** 10.1101/2024.01.10.24301125

**Authors:** David E. Haines, Melissa H. Kong, Peter Ruppersberg, Steven Castellano, Stefan G. Spitzer, Georg Noelker, Andreas Rillig, Tamas Szili-Torok

**Affiliations:** William Beaumont University Hospital, Oakland University William Beaumont School of Medicine, Royal Oak, Michigan, USA; Ablacon Inc., Wheat Ridge, Colorado, USA; Praxisklinik Herz und Gefäße Dresden, Akademische Lehrpraxisklinik der TU Dresden, Dresden, and Brandenburg University of Technology Cottbus-Senftenberg, Institute of Medical Technology, Germany; Department of Cardiology, Heart and Diabetes Center North Rhine-Westphalia, Ruhr University Bochum, Bad Oeynhausen, Germany; University Heart Center, Hamburg GmbH; Department of Cardiology, Erasmus MC, Rotterdam, the Netherlands

**Author notes:** **Corresponding author:** David E. Haines, MD Director, Heart Rhythm Center Professor, Cardiovascular Medicine Oakland University William Beaumont School of Medicine 3601 West 13 Mile Road Royal Oak, MI 48073. **Conflicts of interest:**. DH: Ablacon consultant, equity. MK: Ablacon employee, equity and patent holder. PR: Ablacon employee, equity and patent holder. SC: Ablacon employee, equity. SS, GN, AR, TS: Nothing to disclose.

**Keywords:** Persistent atrial fibrillation, electrographic flow mapping, basket catheter, panoramic mapping

## Abstract

**Background:** Electrographic flow (EGF) mapping algorithms employing Horn-Schunck flow estimations can create temporospatial visualizations of atrial electrical wavefront propagations during atrial fibrillation (AF). Reproducible patterns of centrifugal EGF activation from discrete sites may represent sites of AF origin or sources. Our objectives were to assess the patterns and prevalence of AF sources using EGF mapping.

**Methods:** Unipolar electrograms were recorded for 1-minute with 64-pole basket catheters. Flow estimates were constructed by passing consecutive frames through an algorithm to learn and then compare typical wave direction patterns to describe flow-field evolution. During each 2-second segment, sites initiating centrifugal activation patterns were defined as AF sources. Maps of source location/activity duration were generated.

**Results:** The EGF method was applied to 405 prospective and retrospective patients with persistent or long-standing persistent AF. Mean age 62.5 years; mean LA size 54 mm; mean AF duration 4.6 years. EGF mapping found 6.6 ± 2.4 AF sources/patient (range 1 to 17). Distribution was 55% LA and 45% RA. Dominant sources (prevalence ≥20%) were demonstrated in 185 (45.7%) patients, but only 10.7% of all sources were dominant. While AF cycle length (CL) was not affected by source prevalence, CL variance significantly decreased as source prevalence increased.

**Conclusions:** Complex AF conduction patterns make ablation challenging, but EGF mapping enables detection and organization of time-dependent AF behaviors. Although many low prevalence sources are detected, they may not be clinically relevant, while higher prevalence sources seem to modulate AF. Recording durations of 1 minute facilitate source discrimination.

## INTRODUCTION

Atrial fibrillation (AF) is characterized by chaotic and complex atrial activations that confound clinical attempts at mapping. While the most widely accepted mechanism of AF initiation remains rapid, intermittent focal firing from pulmonary vein (PV) triggers, considerable debate continues regarding the mechanisms of AF initiation and propagation beyond the PVs.^1,2^ Evidence that self-sustaining extra-PV drivers and/or triggers contribute to AF recurrence after pulmonary vein isolation (PVI) continues to expand.^3–5^ Whether these represent sites of focal firing,^1^ microreentry ,^6^ or epicardial to endocardial breakthrough due to longitudinal dissociation of epicardial and endocardial conduction^7,8^ continue to be questions for active debate.

The highly complex wave dynamics of AF continue to complicate efforts to understand the underlying mechanisms responsible for its initiation and perpetuation.^9^ As a result, numerous attempts have been made to localize these sites after PVI^10–15^ but have shown little benefit because of multiple technical limitations. Electrographic flow (EGF) mapping is an innovative method employing Horn-Schunck flow estimations to create temporospatial visualizations of broad atrial electrical wavefronts propagation during AF.^16,17^ Enhanced by innovative machine-learning algorithms, putative sites of origin of AF activation, or “sources” may be identified by reproducible patterns of centrifugal EGF activation from those sites. The purpose of this study was to use EGF mapping to retrospectively and prospectively analyze global left (LA) and right atrial (RA) maps acquired during AF mapping and ablation procedures to assess the prevalence and distribution of source activity in these patients.

## METHODS

### Study population and procedure

Patients with persistent or long-standing persistent AF undergoing AF ablation for clinical indications at 2 different centers (Erasmus Medical Center in Rotterdam, Netherlands; Praxisklinik in Dresden, Germany) were included in this analysis (NCT04473963). Studies were approved by the institutional review board at each institution. After informed consent, all procedures were performed under sedation and systemic anticoagulation was achieved using intravenous heparin titrated to maintain an activated clotting time > 300 seconds prior to insertion of the mapping catheter.

Transseptal catheterization was performed in standard fashion and 3-dimensional electroanatomic maps of both atria were created using Carto (Biosense Webster, Diamond Bar, CA) or EnSite Precision (Abbott, Abbott Park, IL) mapping systems. Recordings from 64-pole basket electrodes (FIRMap™, Abbott, Abbott Park, IL) positioned in the RA and LA were prospectively gathered. Basket electrode recordings were acquired before ablation although 62% of patients had undergone prior AF ablation procedures, then post-processed using EGF mapping algorithms. A second retrospective EGF mapping data set was derived from 64-electrode basket catheter recordings acquired during previous AF ablation procedures from 5 centers (Erasmus Medical Center in Rotterdam, Netherlands; Praxisklinik in Dresden, Germany; Ruhr University in Bad Oyenhausen, Germany; Asklepios Clinic St. Georg in Hamburg, Germany; Charité, Benjamin Franklin Clinic, Berlin, Germany). These data were deidentified and retrospectively analyzed after approval by each of the local institutional review boards.

### Electrographic flow mapping

Using proprietary software (Ablamap^®^, Ablacon, Inc,, Wheat Ridge, CO), EGF maps were generated from the raw unipolar intracardiac EGMs recorded from a 64-electrode basket catheter over 1-minute intervals sequentially in the RA and LA. The details of electrogram acquisition and processing have been described in detail.^18^ In brief, signals were filtered at 0.05 – 500 Hz and recorded at 1kHz sampling frequency for export from the electrophysiology recording system. The EGF mapping system pre-processes unipolar EGMs to remove the time segment containing the QRS complex from the recording and then normalizes the signals to unitary amplitudes prior to processing. The EGF mapping algorithm provides snapshots of the electrical field taken every 19 milliseconds and then collated to determine spatial voltage gradients versus temporal voltage gradients derived from each 2 subsequent frames. Changes in position of voltages and their associated spatial gradients over time create a flow vector matrix displaying excitation waves’ behavior and propagation over time.

### AF Source detection and prevalence

The EGF mapping algorithms create a temporospatial visualization of atrial electrical wavefront propagation during AF over sequential 2-second time segments for the 60-second total mapping acquisition. Dominant patterns of excitation wave propagation are displayed in the flow-vector maps and some of these patterns are repetitive and consistent over time. Sites from which flow vectors reproducibly emerge in divergent patterns (where the continuously existing flow vector directionality has a mathematical pole) are designated as “sources” of electrographic flow. **Figure 1** shows how sources are registered and summation of the results frame-by-frame results in exposure of higher prevalence sites. Summing up the source activity from 105 frames of data for each 2-second segment yields a prevalence of each source at its location for each segment from 0-100%. If a source is detected in all 105 frames in a 2-second segment, the source prevalence or detection rate for that segment is 100%. The prevalence of activity from each source over the 60-second recording is reported in the corresponding prevalence histogram plot that displays cumulative data from each 2-second segment of the full minute (**Figure 2**). Summing the data from all thirty 2-second segments yields the source prevalence over the 1-minute acquisition period. Dominant sources were defined as those sites with source activity occurring ≥20% of the time during the 60 sec signal acquisition.

**Figure 1:**
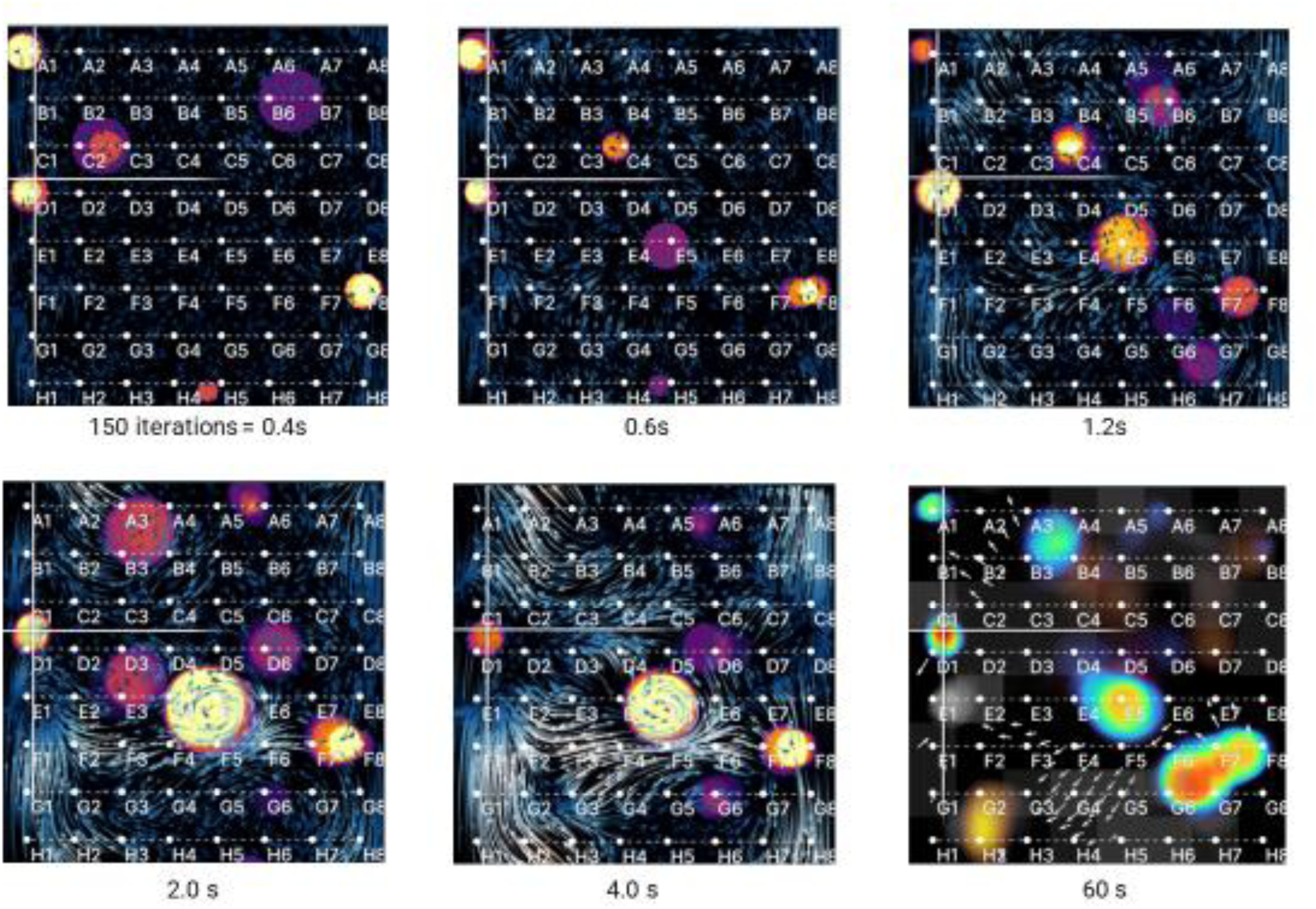
An evolving EGF map showing accruing Summary Map imagery over time is portrayed in a planar view of the basket electrograms. Prevalence values of source activity become increasingly consistent and reproducible over time as an increasing number of frames accumulate over 60 seconds. After 0.4 seconds of recording, the flow-field appears undefined with multiple possible sources estimated (orange spots); however, after 4 seconds of Horn-Schunck iterations, a distinct pattern of two separate sources becomes apparent. After recording for 1 minute, these two distinct sources remain stable and consistent as displayed in the EGF Summary Map showing a “heat map” of the source activity and locations (red spots).

**Figure 2:**
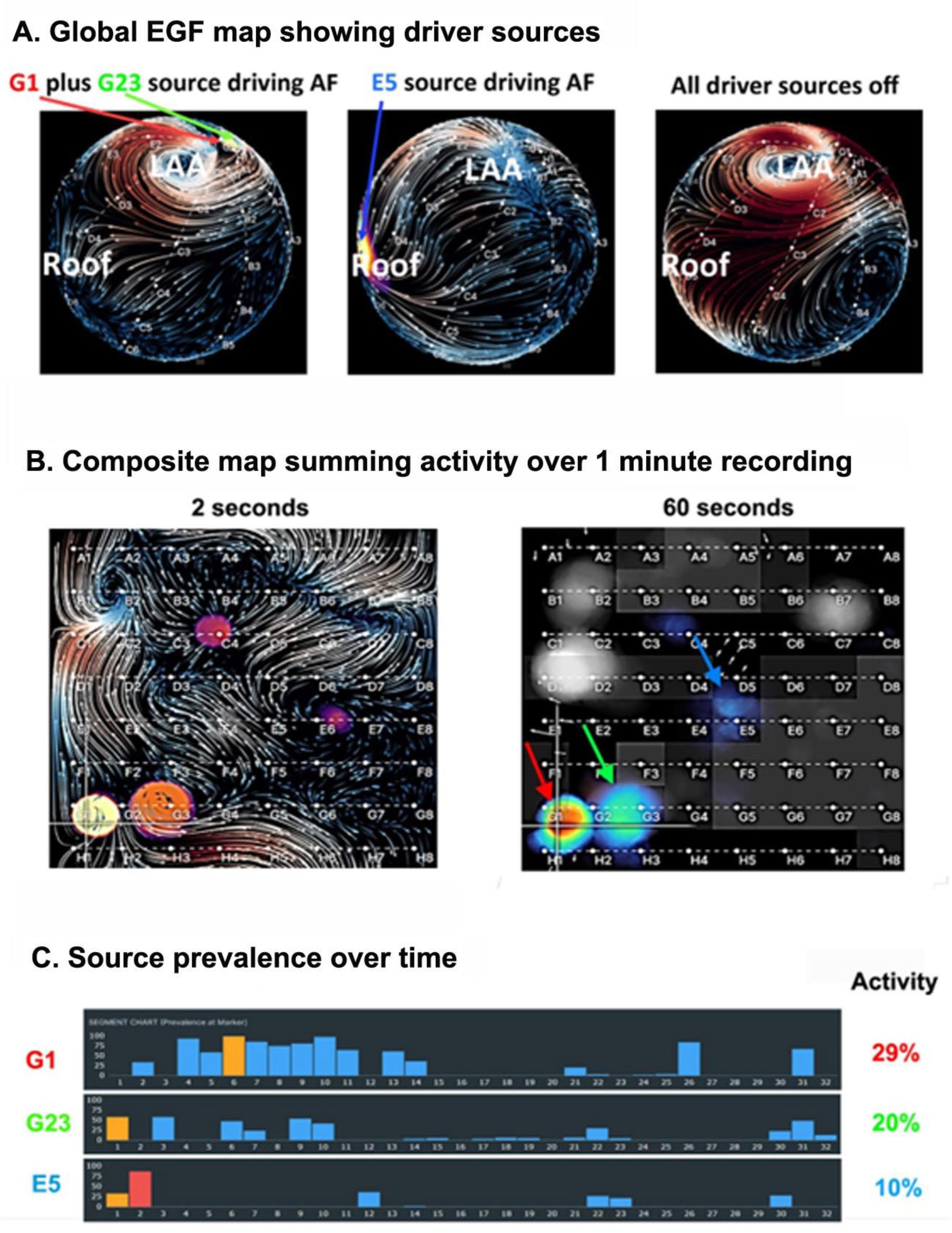
**A.** 3D EGF Segment Maps recorded from central LA basket position and displayed in a global view shows an active source at G1 and G23 during a 2-second segment (left); a separate third source at E5 during another segment (middle); and a segment during which all 3 sources are inactive (right). Note the different flow-patterns present depending on which source(s) are active. **B**. The EGF Segment Map (left) shows the flow-patterns during a 2-second segment. The EGF Summary Map (right) displays source prevalence. **C.** Histogram plots show the time-dependence of source prevalence over the full minute of recording with the most relevant 2-second segments (yellow). The source at G1 has an activity level of 29% while both the source at G23 and E5 have lower activity levels and thus, G1 represents the leading source.

During each 2-second segment, centrifugal or divergent activation patterns define active sources that may represent AF drivers and/or triggers. Passive rotational phenomena are characterized by centripetal or convergent activation patterns as previously described.^16^ All active sources and passive phenomena detected over 1-minute of recording are displayed in the EGF Summary Map (**Figure 2b)**.

Active sources with higher prevalence are red, and those with lower prevalence are blue. Likewise, passive phenomena with higher prevalence appear white and lower prevalence appear grey. Dominant sources are empirically defined as those observed during ≥20% of the recording.

### Cycle length analysis

Cycle length (CL) analysis was conducted on the first recordings with valid collection parameters (sufficient recording time of >55 seconds and no baseline drift) for all prospective patients. CL was computed per FF interval at each electrode location by measuring peak autocorrelation per channel and filtering out electrode locations where autocorrelation was < 0.6. CL per FF interval. These data were then recomputed as CL per 2second segment by binning and averaging the FF intervals by segment at each electrode. If the ratio of near field atrial electrogram amplitude to far-field QRST amplitude was <0.7, those recordings were defined as low contact and were censored from the analysis. The final CLs per 2-second segment were computed by averaging across all remaining electrodes with good atrial wall contact.

For each 2-second segment of EGF recording, the mean AF CL from all electrodes with good contact and the standard deviation (SD) of those values were compared to the prevalence of source activity in the same time segment. The SD values represented the variance in AF CL over all electrode locations over the 2 second recording time segment and were interpreted as the variance in beat-to-beat FF interval across the basket electrode’s recording area in the atrial chamber. The prevalence of source activity (0.0 to 1.0) was divided into deciles, and the data were presented in bins representing mean and SD values for each decile.

### Statistical analysis

Continuous variables are summarized as mean value ± one standard deviation. Categorical variables are shown as percentages. Two-tailed, two sample t-tests were used to assess the difference between the independent means of continuous variables between subgroups. Fisher’s exact test was used to compare proportions between subgroups. Two-tailed, paired t-tests were used to assess differences in CL at low versus high source prevalence. Linear regression analysis was used to assess AF CL and AF CL SD trends. For all statistical tests, the null hypothesis was rejected at the level of p < 0.05. Lead authors (D.H., M.K., P.R. and S.C.) had full access to all the data in the study and take responsibility for its integrity and the data analysis.

## RESULTS

### Patient population

The pooled study cohort included data from 399 patients with persistent or long-standing persistent AF who underwent 64-electrode basket recordings of intracardiac EGMs either as part of a FIRM-guided or a prospective EGF-guided AF ablation procedure. For patients who underwent a FIRM-guided procedure, the deidentified raw unipolar recordings were retrospectively analyzed and processed post-hoc. For patients who underwent a prospective EGF-guided procedure, the EGF maps were recorded and generated in near real-time. The study cohort’s mean age was 63.4±9.2 years and 41% were female. Mean AF duration prior to mapping was 51±42 months; 62% had undergone prior PVI and 63% had been treated with at least one class I or III antiarrhythmic drug. Mean short axis left atrial dimension was 50±8 mm. The mean CHA_2_DS_2_-VASc-score was 2.2±1.4. Additional baseline patient demographics including comorbidities are shown in **Table 1**.

**Table 1:** Baseline Demographics.

|  | Total | No dominant source<br>(temporal prevalence<br>< 20%) | Dominant source present<br>(temporal prevalence<br>≥20%) | P value |
| --- | --- | --- | --- | --- |
| Number of patients, n | 399† | 217† | 182† |  |
| Age (years) (mean, SD) | 63.4±9.2 | 62.4±8.8 | 64.6±9.6 | 0.005 |
| Sex (Female), n (%) | 132 (41.4) | 58(33.7) | 74(50.3) | 0.003 |
| Body mass index, (mean, SD) | 29.1 ± 5.2 | 29.6 ± 5.6 | 28.6 ± 4.6 | 0.23 |
| Left ventricle ejection fraction<br>(mean, SD) | 0.57 ± 0.08 | 0.57 ± 0.08 | 0.57 ± 0.08 | 0.77 |
| Left atrial size (mm) (mean, SD) | 50.5 ± 7.5 | 51.8 ± 7.2 | 48.8 ± 7.4 | 0.001 |
| AF duration (months) (mean, SD) | 51.3 ± 41.5 | 53.9 ± 42.8 | 48.2 ± 39.9 | 0.23 |
| Prior AF ablation, n (%) | 194 (62.2) | 120(70.6) | 74(51.7) | 0.001 |
| Prior antiarrhythmic drug use, n (%) | 168 (63.4) | 91(60.7) | 77(67.0) | 0.08 |
| Hypertension, n (%) | 204 (73.6) | 116(77.9) | 88(68.8) | 0.10 |
| Diabetes mellitus, n (%) | 45 (18.5) | 26(18.8) | 19(18.1) | 1.0 |
| History of CVA/TIA, n (%) | 22 (9.1) | 12(8.8) | 10(9.3) | 1.0 |
| Coronary or vascular disease, n (%) | 52 (28.1) | 27(25.0) | 25(32.5) | 0.32 |
| CHA<sub>2</sub>DS<sub>2</sub>-VASc-score‡ (mean, SD) | 2.2 ± 1.4 | 2.0 ± 1.3 | 2.5 ± 1.5 | 0.004 |

†Total number of patients analyzed for source activity level; however, for each of these patients, not all baseline demographic variables were available.
‡CHA<sub>2</sub>DS<sub>2</sub>-VASc-score is a calculated risk stratification score to predict risk of stroke in AF patients.
AF: atrial fibrillation; CVA/TIA: cerebrovascular accident/transient ischemic attack; SD: standard deviation

### Prevalence of detected sources

Among the 399 prospective and retrospective patients with persistent or long-standing persistent AF in this study, a total of 2,625 sources with prevalence > 5% were detected, ranging from 1 to 17 per patient with a mean of 6.6 ± 2.4 sources per patient (**Figure 3**). Sources with very low prevalence (<5%) were not included. Of the 2.623 sources detected, over half of all sources detected had a low prevalence of 5-10%. Only 278 sources (11%) from 182 patients (46%) met criteria for dominant sources with a prevalence ≥20%. The dominant sources were distributed between the LA (55%) and the RA (45%). Patients with a dominant source were older (p=0.005), more likely to be female (p=0.003), had smaller LA size (p<0.001), and had a greater likelihood of previous AF ablation procedures (p=0.001) compared with those without dominant sources (**Table 1**).

**Figure 3:**
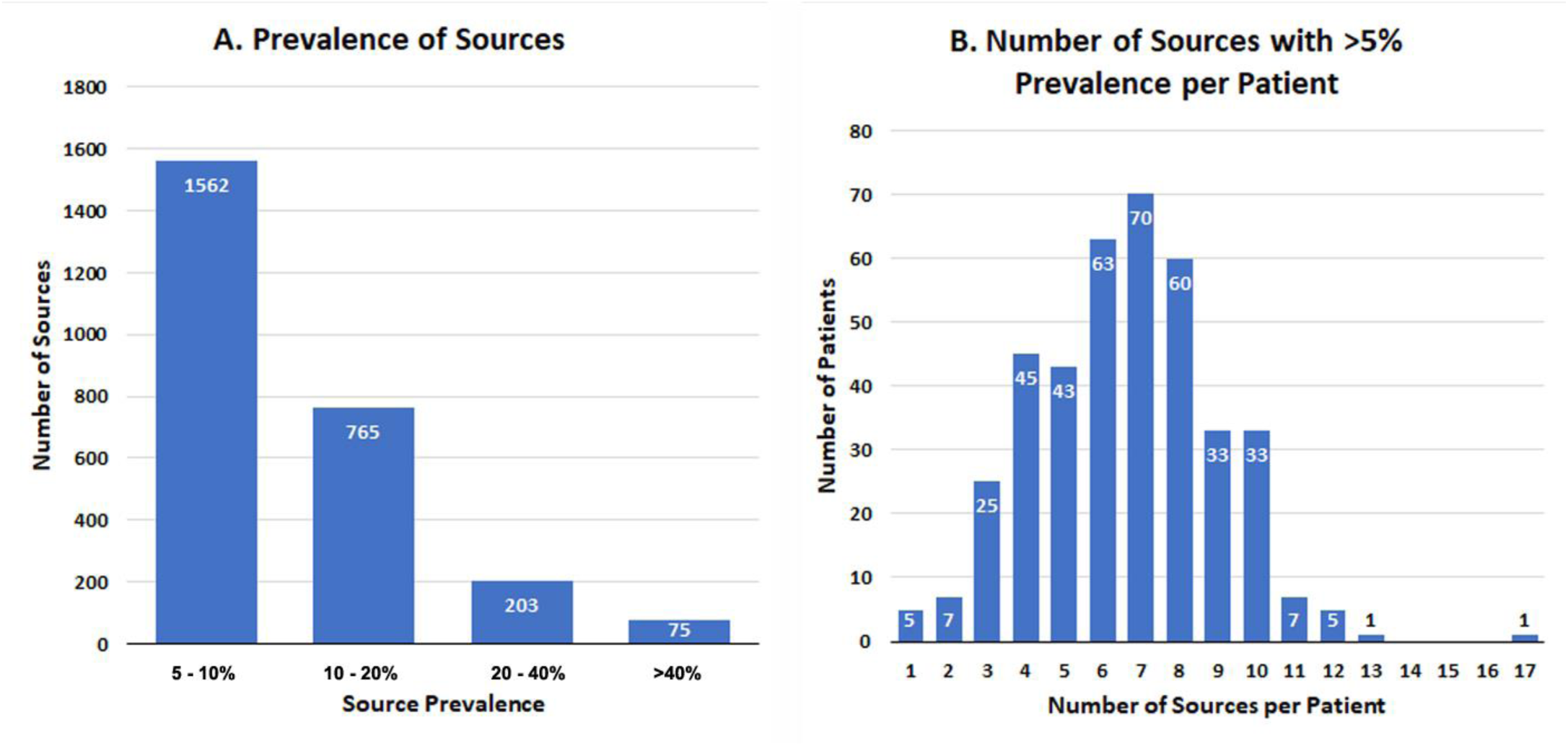
**A.** A histogram of the prevalence of all sources over a 60-sec mapping acquisition is displayed. Only 11% of sources are dominant sources with prevalence ≥20%. **B.** The number of sources with > 5% prevalence per patient ranged from 1-15 is shown in this histogram.

### Behaviors of sources

During EGM recording, sources were dynamic and would alternate between activity and inactivity. This time-dependent behavior of AF is demonstrated in the patient example shown in **Figure 4**. In this example, there were 3 distinct sources of AF, each located in a different part of the LA and each demonstrating a different local pattern of activation when the source was active. When more than one source activated, a local region of influence from the active source or sources was observed based on the local activation patterns. When two sources were simultaneously active, Flow Origin Maps manifested the regions of influence of each source. Although sources switched off and on and demonstrated different prevalences (percentages of the time during which they were activated), these origins of excitation did not appear to wander.

**Figure 4:**
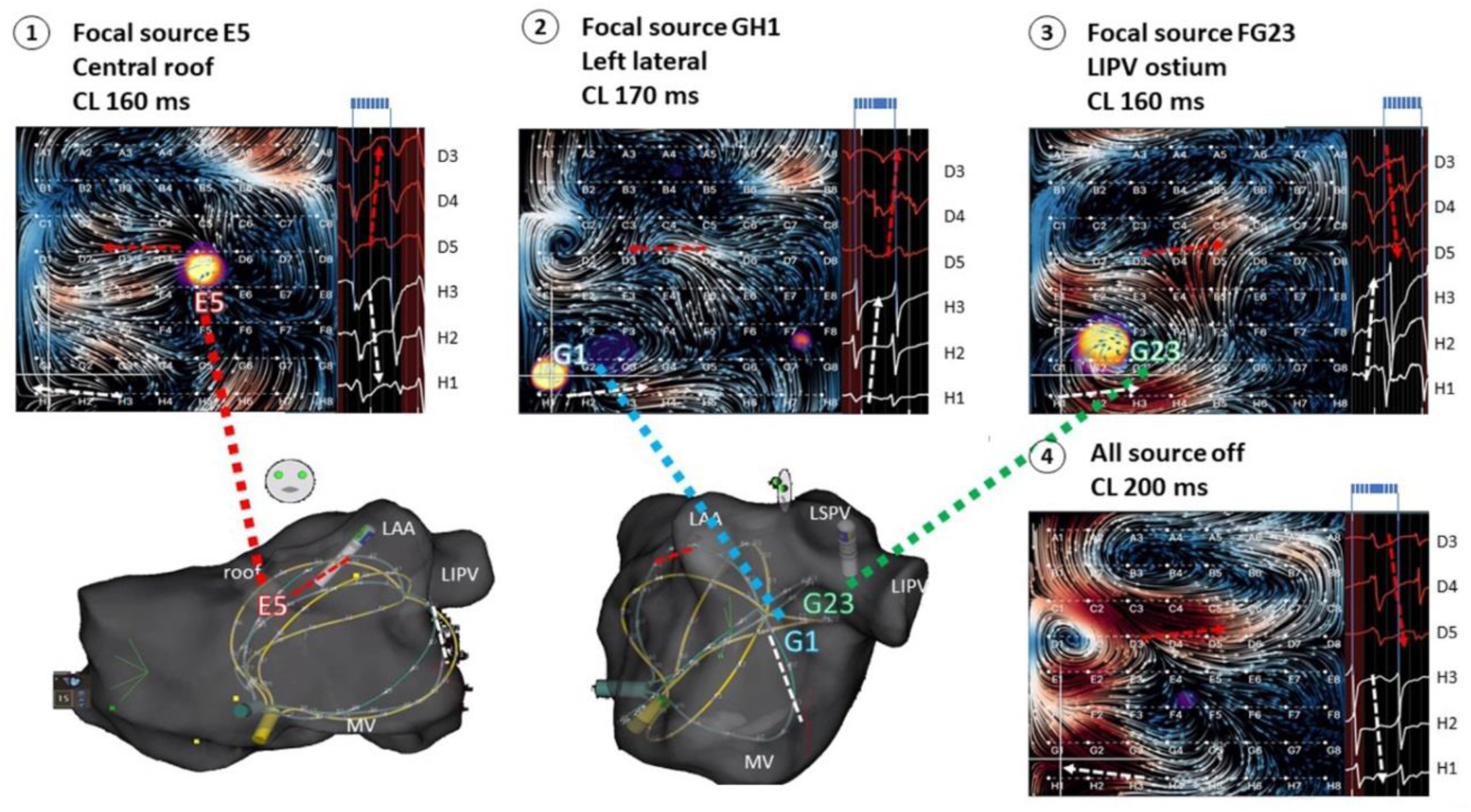
Individual 2-second segments of the 60-second EGF acquisition in this patient example demonstrated different EGF patterns of AF activation depending upon the activity of the 3 independent sources. Two-second EGF maps show focal source activity on the LA central roof (panel 1), at a left lateral site (panel 2), and near the LIPV ostium (Panel 3). Panel 4 shows no source activity. The electroanatomical geometries of the basket electrode catheter positioned in the left atrium are displayed in AP (left) and left lateral (right) projections.

### Effect of source activity on AF cycle length

AF CL was not affected by prevalence of source activity. The mean AF CL across all patients during 2-second periods of zero source activity was 222 ± 10 ms compared to 221 ± 7 ms during 2-second periods of 100% source activity (**Figure 5a**, p = 0.320). Therefore, source activation did not cause acceleration of AF in surrounding passively activated atrium. However, entrainment of surrounding atrium by activation originating from the source could be imputed by the observation that CL variance significantly decreased as source prevalence increased. The CL standard deviation during zero source activity was 10.4 ms versus 6.5 ms when the source was continuously activated (p < 0.001). A significant trend of decreasing AF cycle length variance was observed as prevalence of source activity increased during the 2-second acquisition windows (**Figure 5b**, p < 0.001).

**Figure 5:**
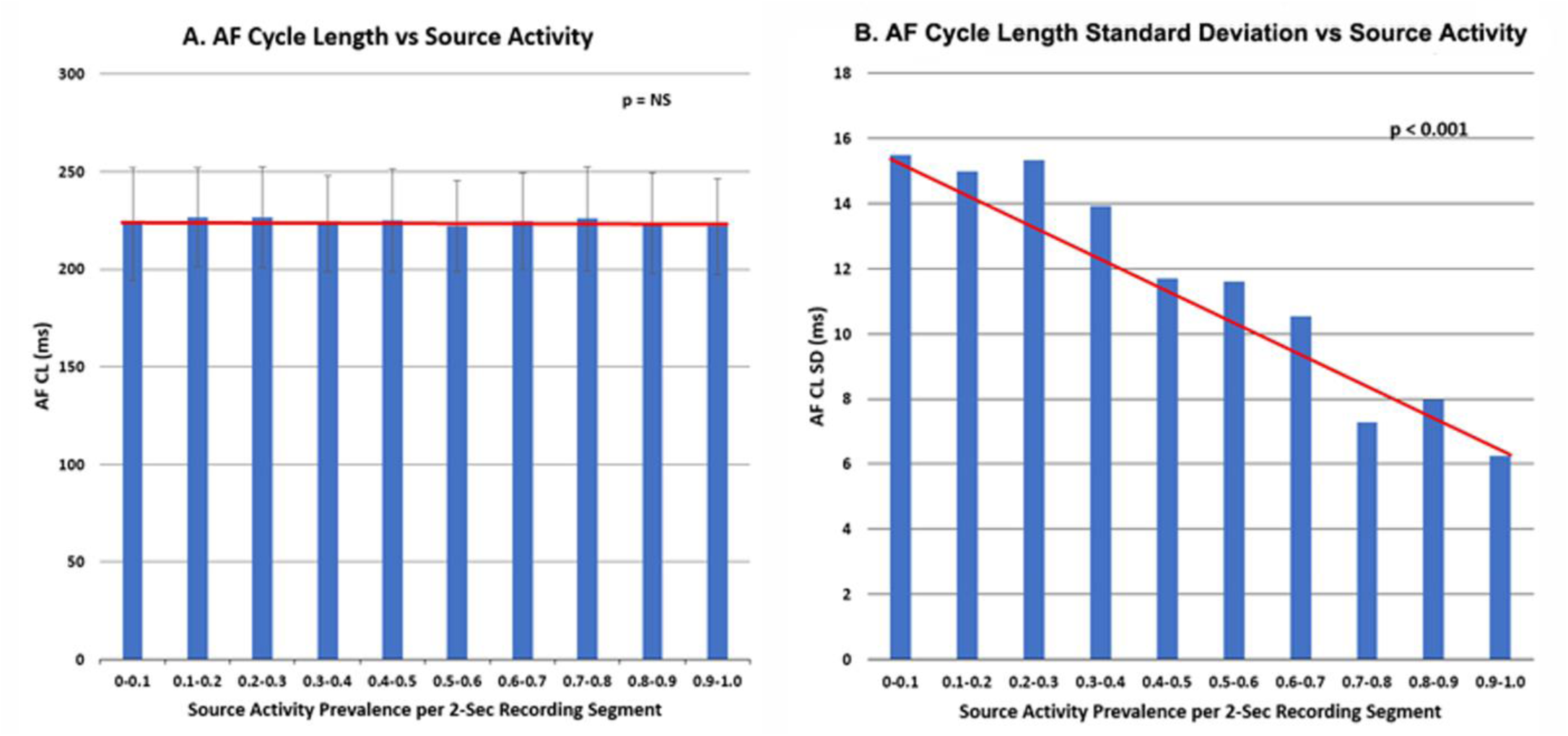
**A.** The AF CL versus source activity prevalence per 2-second recording segment is shown by decile. Little effect on the AF CL of the recording regions surrounding the putative AF source is seen. The linear regression line for the individual data is superimposed (p = NS). **B.** The variance of the AF CL the surrounding regions represented by the standard deviation versus the prevalence of source activity for each 2-second segment is shown. With increasing source activity (presented as average standard deviation per decile of source activation within the 2-second segment), there is significant reduction in AF CL variance indicating that the AF source is entraining the surrounding atrial recording sites. AF CL = atrial fibrillation cycle length. SD = standard deviation.

## DISCUSSION

PVI has become a widely accepted and effective treatment strategy for patients with symptomatic, drug-refractory paroxysmal AF.^19^ Despite favorable long-term clinical success rates of PVI and its establishment as the cornerstone of non-pharmacologic AF treatment, catheter ablation for persistent AF remains hampered by suboptimal long-term success rates.^20,21^ Although debate still surrounds the underlying pathophysiology of persistent AF, a growing body of data supports the role of self-sustaining, localized extra-PV AF drivers and triggers.^5,22,23^ These focal activations have been documented in close proximity to passive rotational phenomena as well.^13,24^ As a result, a wide array of strategies for the detection and elimination of clinically relevant extra-PV ablation targets have evolved. However, studies employing these various mapping techniques have yielded inconsistent clinical outcomes.^10,11,13,14,25,26^

The present study employed a novel mapping algorithm using atrial electrograms recorded from a low-density 64-pole basket, and Green’s formula-based spline interpolation with Horn-Schunck’s iterative flow estimation to generate reproducible patterns of electrographic flow.^18^ Although the present system is subject to the same limitations experienced by other systems that have employed low-density basket electrode recordings, our goal is to identify broad wavefronts of activation, not to dissect the intricacies of source mechanisms at their sites of origin. The anticipated wavelength of these broad atrial electrical wavefronts is estimated to be greater than 10 cm, which is significantly greater than the interelectrode spacing of a basket electrode catheter and therefore within the resolution of the low-density electrode array. In this study, EGF mapping was able to identify discrete atrial anatomical sites that showed reproducible patterns of centrifugal activation from putative sites of origin for AF initiation and propagation. AF source prevalence ≥5% was 6.6 per patient, and 11% of those sources had ≥20% prevalence. Although duration of activation of these sources was often <2 seconds at a time, the pattern of recurrent activation originating from discrete sources was observed in all patients. A previous report demonstrated the reproducibility and spatiotemporal stability of the EGF-identified sources of AF over long time periods in an individual patient.^27^ It is hypothesized that substrate modification at sites of higher source activity should have a salutary effect on prevention of clinical AF. It is also hypothesized that the pattern of EGF activity and prevalence of source activity will be useful in the phenotyping of subtypes of clinical AF and may help determine which patients are the most likely to respond to catheter ablation or not.

Source activation was not associated with shorter AF CL in surrounding passively activated myocardium indicating that source activity is not necessarily faster than surrounding fibrillatory conduction. There may be more wave front collisions with complex electrograms and short local cycle lengths when sources are dormant, and then more coordinated, entrained conduction when sources activate. What was observed, however, is that greater prevalence of source activity resulted in significant reduction in surrounding cycle length variability consistent with regional and remote entrainment from the active source.

### Alternative mapping strategies for identifying sites of AF initiation and propagation

The disappointing clinical results of a range of ablation strategies to eliminate persistent and long-standing persistent AF is likely rooted in the challenge of identifying the critical substrate required for initiation and propagation of this arrhythmia. To improve these outcomes, a variety of mapping strategies have been proposed. EGM-based quantitative approaches for targeting locations with maximal dominant frequency or complex fractionated atrial EGMs (CFAEs) have been proposed.^3,28–32^ These nominal signal analyses require time-intensive offline analysis, yield inconsistent results, ^3,9,28,33–35^ are sensitive to errors related to timing, signal morphology, noise, and EGM amplitude, and lack the ability to estimate time-dependent electrical activity.^36^

Other algorithm-based approaches developed for the detection of extra-PV AF sources have been based on phase-mapping or activation mapping.^10,11,13,25,26^ Phase-analysis approximates a periodic function, which results in the creation of artifacts, false positives and epiphenomena when applied to the chaotic process with low-amplitude EGMs observed with AF.^37^ The inherent tendency to assume a periodic process and rotational structure results in the inability to identify focal activations with divergent wavefront propagation origins without phase singularities; and the inability to extrapolate excitation wave propagation vectors from estimated phase angles renders phase-analysis unable to distinguish between active AF drivers and passive bystander rotational phenomena.^24^ Phase maps also suffer severe limitations regarding the detection of time-dependent relevance or stability over longer recordings. Technologies based on activation mapping suffer from noise sensitivity due to the need for accurate assignment of local activation time during AF. Mapping technologies that utilize regional or stamp mapping rather than global or panoramic mapping techniques cannot see the inherent high variability of the electrical flow-fields in AF across the spectrum of time. Beyond the limited detection of time-dependent electrical behaviors, none of the above methods offer real-time or beat-by-beat mapping and largely require time-intensive offline signal processing.^11,26,38,39^

### EGF mapping detects and summarizes time-dependent AF behaviors

Although numerous methodologies have been developed to identify potentially effective AF ablation targets beyond the PVs, given the above-described limitations of available mapping technologies and algorithms, accurate and effective mapping of AF presents many technical challenges. EGF mapping may offer a solution for surmounting many of these problems. EGF mapping does not assume periodic activity and is not preprogrammed to look for any specific phenomena, but instead generates near real-time visualizations of global, atrial wavefront propagation at any given basket electrode position. Since a 60 second acquisition has been selected as an appropriate duration to provide good sensitivity for source identification and good reproducibility (ref), acquisitions from overlapping basket positions can provide a complete biatrial map in minutes.^40^ Using an optical flow algorithm that triangulates between electrodes (assuming electrode distance exceeds excitation wavelength) provides a spatial resolution beyond the interelectrode distances of the multielectrode basket catheter. Unipolar recordings from the endocardial surface of the atria naturally favor the display of endocardial electrographic flow. However, it is likely that EGF mapping incorporates some epicardial signal and partially represents both endocardial and epicardial flow. Assessing electrographic flow to identify sources rather than attempting to map the precise activation mechanism of the source (e.g. focal activation, microreentry, or epicardial breakthrough) provides actionable data with high temporal resolution despite the use of low-resolution mapping. Enabling the global mapping of the atria provides efficient visualization of time-dependent electrical behaviors.

EGF mapping is a promising approach that enables the creation of near real-time temporospatial visualizations of atrial electrical wavefront propagation to identify putative AF sources. Although the electrical flow-fields during AF are inherently variable and seemingly chaotic, the origins of EGF— defined as sources, often recur in the same locations. As such, because EGF Summary Maps display the dominant patterns of excitation wave propagation from the resulting flow-vector maps generated for each 2-second segment, these EGF Summary Maps can organize these seemingly stochastic flow-fields by integrating this repetitive behavior over the time domain. Ongoing research and development of the EGF mapping algorithms should enable the separation of endocardial and epicardial flow, particularly if they are dyssnchronous over time, i.e., if they occur at different time points in the recording due to differences in refractory periods of different layers of myocardium.

Additionally, EGF mapping can be used to accurately identify simulated sources of AF during spontaneously persistent AF in an animal model^41^ and appears to produce consistent and reproducible intra- and inter-procedural maps in humans.^40^ A first-in-human case report presented by Szili-Torok et. al, recently demonstrated inter-procedural EGF map reproducibility with spatiotemporal stability of EGF-identified sources of AF in the same patient during procedures performed 18 months apart.^27^ In a double-blinded retrospective study of 64 patients, EGF-identified sources with high activity (>26%) appear to be clinically relevant and their presence post-ablation correlated with high rates of AF recurrence.^42^ More recently, a single-center, prospective study of 70 patients with persistent AF found that among patients who received PVI + EGF-guided source ablation, only 25.6% of patient has AF recurrence at 12-months post-procedure while more than half of the patients having recurrent AF among those who received PVI-only (62.5%) and those who underwent PVI + linear ablation (53.3%), p = 0.02).^43^ The results from the completed randomized controlled clinical trial, *FLOW-AF* (NCT 04473963) that was designed to evaluate the effectiveness of EGF mapping for AF source detection and for targeting ablation therapy in persistent and long-standing persistent AF patients were recently presented and showed that EGF mapping and ablation resulted in successful elimination of sources in 95% of patients randomized to source ablation and that PVI with adjunctive EGF-guided source ablation improved AF free survival by 51% on an absolute basis at 1 year compared with PVI-only.^44,45^

### Study Limitations

Despite the robustness of the EGF mapping signal processing, the limitations of using a basket catheter to record intracardiac signals cannot be ignored. Low signal-to-noise ratios secondary to non-uniform endocardial contact in addition to catheter deformation with spline splaying and bunching and incomplete atrial coverage present potential pitfalls in terms of the quality of the data acquisition for analysis. It should be noted that positive detection of EGF sources with high activity is only possible if the recorded signals accurately indicate such a flow pattern. However, the absence of sources in a single atrial recording of one minute does not necessarily mean that no sources of AF can be detected in the EGF mapping of this atrium. To successfully exclude the presence of a source, multiple positions of the basket catheter in the same atrium should be recorded and analyzed for several minutes each. Similarly, if sources exist only in one position in one of the two atria, it would be necessary to reinvestigate both atria after source ablation to exclude the possibility that another source has taken over as the dominant flow origin upon the elimination of the original source that had been dominating the flow. Finally, all of the recordings analyzed were taken prior to ablation, which might change the source prevalence post-ablation.

### Conclusions

Complex AF conduction patterns make the detection and discrimination of clinically relevant AF sources difficult. The EGF mapping algorithm is uniquely able to utilize low density multielectrode basket EGMs to characterize the time-dependent centrifugal electrical activations reproducibly emanating from discrete sites that have been designated as AF sources. As they are present in more than half of patients with persistent and long-standing persistent AF, modulating these sources with catheter ablation may have a salutary effect on preventing AF recurrence post-ablation and further clinical studies are warranted.

## Data Availability

The raw data analyzed in this study is readily available for review.

## Acknowledgments, Sources of Funding, & Disclosures

**Sources of Funding:** Funding for data collection and analysis were supported in part by the Beaumont Foundation with a generous gift from Kimberly A. Whipple, and from Ablacon, Inc., Wheat Ridge, CO.

**Disclosures:** Ablacon, Inc. equity (D.H., M.K., P.R. and S.C.); Ablacon, Inc. employee ( M.K., P.R. and S.C.)

## Non-standard Abbreviations and Acronyms

AF =: atrial fibrillation
EGF =: electrographic flow

